# Time to Recovery from Diabetic Ketoacidosis and its Predictors among Adult Diabetic Ketoacidosis Patients in DEBRE MARKOS Referral Hospital, North West Ethiopia, 2021: Retrospective Cohort Study

**DOI:** 10.1101/2022.04.12.22273779

**Authors:** Dessie Temesgen, Yihun Miskir, Getenet Dessie, Ahmed Nuru, Berihun Bantie Tesema, Molla Azmeraw, Abraham Teym, Melesew Dagne

**Affiliations:** Department of Nursing, Collage of Health Science, Woldia University, Woldia, Ethiopia; Department of Emergency and Critical care Nursing, School of Health Science, Collage of Medicine and Health science, Bahirdar university, Northwest Ethiopia, Ethiopia; Department of Adult Health Nursing, School of Health Science, Collage of Medicine and Health science, Bahirdar university, Northwest Ethiopia, Ethiopia; Departments of Nursing, Collage of Health Science, Wolkitie University, Wolkitie, Ethiopia; Department of Adult Health Nursing, Collage of Medicine and Health science, Debre Tabor University, North west Ethiopia, Ethiopia; Departments of Environmental Health, Collage of Health Science, Debre Markose University, Debre Markose, Ethiopia

**Keywords:** Diabetic ketoacidosis, recovery time, Debre Markose, Ethiopia

## Abstract

**Introduction:** Diabetic ketoacidosis is an acute life-threatening complication of diabetes mellitus. With appropriate treatments, diabetic ketoacidosis patients are expected to make a full recovery within 24 hours. Previous studies did not address variables such as duration of diabetic ketoacidosis symptoms, and blood glucose level. In addition, the recovery time and its predictors of diabetic ketoacidosis in adult patients are not well known in Ethiopia.

**Objective:** To assess time to recovery from diabetic ketoacidosis and its predictors among adult diabetic ketoacidosis patients in Debre Markos referral hospital, North West Ethiopia, 2021

**Methods:** A retrospective cohort study was employed among 452 records of adult diabetic ketoacidosis patients who were admitted starting from January 1, 2016 to January 1, 2021 using their medical registration number. Data were entered into Epi-data version 4.6 and analyzed using Stata version 14. A Kaplan Meier survival curve was used to estimate diabetic ketoacidosis-free survival time. In addition, a generalized log-rank test was utilized to compare diabetic ketoacidosis-free survival time between different categorical explanatory variables. Cox proportional hazards model was used to identify predictors of time to diabetic ketoacidosis recovery time. Variables with a P-value < 0.25 in the bivariable analysis were entered into a multivariable Cox proportional hazards model to identify predictors of recovery time at p≤ 0.05.

**Result:** The median time to recovery from diabetic ketoacidosis for all observations was 24 hours. Severity of diabetic ketoacidosis (AHR=0.24, 95%CI=0.16-0.35), duration of diabetic ketoacidosis (AHR=0.46, 95%CI 0.33-0.64), diabetes duration (AHR=1.74, 95%CI 1.35-2.25), and random blood sugar level (AHR=0.64, 95%CI= (0.51-0.79) were significant predictors of recovery time.

**Conclusion and recommendation:** The median time to recovery from diabetic ketoacidosis was relatively prolonged. The hospital shall give special attention to patients with the identified predictors. Further study using a prospective design by including admission pH and admission serum potassium level is advised.

## Introduction

Diabetic ketoacidosis (DKA) is the foremost common acute hyperglycemic crisis in individuals with diabetes mellitus (1). DKA occurs due to the consequence of an absolute or relative lack of insulin and concomitant elevation of counter-regulatory hormones, usually resulting in the triad of hyperglycemia, metabolic acidosis, and ketosis (2). If the initial diagnosis of type1diabetes is delayed or if type 2 DM precipitated by major surgery, severe illness such as myocardial infarction or infection usually presents with DKA (3, 4). DKA occurs mostly in young people with uncontrolled type 1 diabetes mellitus (T1DM) but can also occur in adults with poorly controlled type 2 diabetes mellitus (T2DM) (1, 2).

The American Diabetes Association(ADA) classifies DKA by severity depending on the degree of acidosis and altered sensorium as mild (Arterial pH 7.25–7.30 and Alert), moderate (Arterial pH7.00 to<7.24 and Alert/Drowsy), or severe (Arterial pH<7.00 and Stupor) (5). The diagnostic criteria for DKA accepted by most experts in the *fi*eld are a blood glucose(BG) greater than 250 mg/dL, pH lower than 7.3, serum bicarbonate lower than 15 mEq/L, increased anion gap metabolic acidosis >12, and a moderate degree of ketonemia (beta-hydroxybutyrate greater than 3mmol(6, 7).

Episodes of DKA typically require an emergency department visit or hospital admission for the patient to receive insulin, intravenous (IV) *fl*uids, and electrolyte correction (8). The individualization of treatment based on a careful clinical and laboratory assessment is needed including restoration of circulatory volume and tissue perfusion, resolution of hyperglycemia, and correction of electrolyte imbalance and acidosis(5). Most people are treated initially with intravenous insulin and a two-bag method of fluid replacement is often used intravenous fluid without dextrose initially and, upon volume correction, infusion with dextrose correction to prevent hypoglycemia caused by the insulin therapy(6). The management of DKA includes IVs administration of 1-2L of 0.9% NaCl over 1*-*2 h then continues 0.9% NaCl until BG level reached to 200-250 mg/dL then change to 5% dextrose in 0.45% NaCl and an IV bolus of regular human insulin of 0.1 U/kg, followed by a continuous infusion of 0.1 U/kg/h then reduce insulin rate to 0.05 U/kg/h when BG< 250 mg/dL; in addition to these if K+ level is 3.3*-*5 mEq/L, add 20-40 mEq potassium chloride to replacement fluid and if K+ level <3.3 mEq/L hold insulin; give 20-30 mEq/h until > 3.3 (8). It is also important to treat any correctable underlying cause of DKA such as sepsis, myocardial infarction, or stroke (5).

Timely diagnosis, comprehensive clinical and biochemical evaluation, and effective management are key to the successful resolution of DKA(9). Resolution of DKA occurs when blood glucose is *<* 200 mg/dL and 2 of the following have occurred: a serum bicarbonate level ≥ 15 mEq/L, a venous pH *>* 7.3, a calculated anion gap ≤ of 12 mEq/L, and blood ketones <0.6 mmol/L (10, 11). Previous studies show that using standardized protocols for DKA management improves DKA recovery (12, 13).

DKA patients with appropriate treatment are expected to make a full recovery within 24 hours (9, 11, 14, 15). Prolonged recovery of DKA leads to hypoglycemia and hypokalemia (9), renal failure, coma, cerebral edema, cardiac arrhythmia(16, 17), lung edema with respiratory failure, and thromboembolic complications(18), and leads to death (19, 20).

Timely diagnosis and initiation of management of DKA is associated with the recovery and prevents hypoglycemia and hypokalemia (9); renal failure, coma, cerebral edema, and cardiac arrhythmia (16, 18), hyperchloremia metabolic acidosis, hypophosphatemia, and thromboembolic complications (18). This in turn decrease bed occupancy in the hospital, length of hospital stay and reduce the amount of human and material resource needed to control it as well as its complications.

As DM prevalence has reached alarming levels, DKA and its complication are also increasing (4). Globally the incidence of DKA among T1DM adult patients ranges from 0–56 per 1000 person-years (PYs) and eleven studies reported prevalence with a range of 0–128 per 1000 people (21). In Ethiopia, a meta-analysis study reported there was a history of DKA in 19.9% of cases (both T1DM and T2DM) at Dessie Hospital, and in 62.0% of cases (for T1DM) at the Black Lion Hospital in Addis Ababa (22). Another study conducted in Dilla University Referral Hospital on the magnitude of DKA in newly diagnosed patients with T1DM was found to be 38% (23) and in Hawassa university comprehensive specialized hospital was 40%; among them, 28.2% and 11.8% were with T1DM and T2DM respectively(24).

DKA is the main reason for the hospitalization of patients with DM (25) and its treatment represents a substantial economic burden in terms of health care costs, both directly and indirectly, ranging from individual to national economy. For example in United Kingdom (UK), the average cost for an episode of diabetic ketoacidosis was *£*2064 per patient(26) and in the United States of America (USA), the total charges among DKA patients in 2017 were $6,757,748,178, with a mean of $30,836.19(27).

Even though DKA patients with appropriate treatment are expected to make a full recovery within 24 hours (9, 11, 14, 15) the resolution time of DKA is longer in African countries. For example in South Africa the average time to resolution of DKA was 21 hours; excluding severe DKA, mild and moderate DKA had an average time to resolution of 20 hours (28) and in Kenya, the median time to resolution was 59 hours(29).

The study conducted in Australia reveals that independent predictors of a slower time to resolution of DKA are lower admission pH levels and higher admission potassium levels (30). The previous study recommends that regular review of patients’ biochemical parameters and strict implementation of hospital protocols may reduce complications and shortens the recovery time of DKA (30).

The previous study did not address the rates and quantity of potassium supplementation prescribed, rates of other adverse events such as hypoglycemia, the duration of DKA, blood and glucose level. In addition, as far as my literature search, there was no study in Ethiopia reporting the estimated survival period of recovery from DKA. Therefore, this study will estimate the median time to recovery from DKA and its predictors among adult patients admitted to the Debre Markos referral hospital in Ethiopia.

## Methods and Materials

### Study design

An institution-based retrospective cohort study design was employed to assess time to recovery from DKA.

### Study area and period

This study was conducted at Debre Markos referral hospital which is located in the city of Debre Markos 265 Km from Bahir-Dar and 300 kilometers from Addis Ababa. The hospital is the only referral hospital in the East Gojjam zone and serves around 3.5 million people and 100 health centers and 9 district hospitals are available in the catchment area of the referral hospital (44). According to information obtained from the administrative office of the hospital, around 348 DKA patients are admitted per year. The study was conducted from January 1, 2016, to January 1, 2021, among adults who were admitted with DKA, and registration charts were retrieved *from* March 18-April 18, 2021.

### Population

#### Source population

All adult DKA patients admitted to Debre Markos referral hospital

#### Study population

Adult DKA patient admitted to Debre Markos referral hospital during the study period and whose charts were available

### Eligibility criteria

#### Inclusion criteria

Adult DKA patients above 15 years old and admitted to the hospital with DKA

#### Exclusion criteria

Adult DKA patients who were admitted to the hospital with incomplete and unavailable medical records

### Sample size determination and procedure

#### Sample size determination

To determine sample size predictors significantly associated with the outcome variable were considered. Accordingly, the sample size was determined using a double population proportion formula by considering the duration of DM and presence of CKD as predictor variables of DKA recovery based on a study done in Saudi Arabia (33) by using Epi -info version 7 statistical package. It is calculated by taking a two-sided significant level (α) of *5* %, 95% confidence level, power 80 %, and 1:1 ratio of non-exposed to expose. Finally 464 were selected as the final sample size for the study.

By adding 10% missing data then finally, the largest sample size (N= 464) will be selected as the final sample size for the study.

#### Sampling procedure

The records of adults who were admitted with DKA in the hospital from January 1 2016 to January 1 2021 were selected using MRN. The patient’s charts were selected using a simple random sampling technique by a computer-generating method. Finally, data were extracted from the selected medical charts.

#### Variable of the study

The outcome variable for this study is time to recovery from DKA. The independent variables in this study are age, sex, residence, admission PH, admission potassium, blood urea, serum creatinine, hemoglobin level, initial blood glucose level, urine ketone, severity of DKA, preceding infection of DKA, presence of Comorbidities, duration of DM, vital sign at presentation, frequency of DM follow up, treatment regimen, amount of IV fluid, potassium supplement, initiation time of management and glucocorticoid medication use.

### Operational definition

#### Event

recovery from DKA during the study period

#### Censored

Referred, died, or discharged cases for any reason before recovery from DKA during the study period.

DKA: Adults admitted with blood glucose greater than 250 mg/dL, pH lower than 7.3, and a moderate degree of ketonemia (>3mmol)/positive urine ketone (6, 7).

DKA recovery time: the interval in hours between the first vascular filling to treat the DKA in the emergency room or the ICU to DKA resolution

DKA Resolution: the occurrence of the first blood ketone < 0.5 mmol/l, or the second negative ketonuria (1 or 2 crosses) (32).

Prolonged DKA recovery: the resolution time of DKA is longer than 24 hours (9, 11, 14,15).

#### Incomplete data

when the ketone negative tests and initiation time of management are not recorded.

#### Entry date

the time of admission with DKA diagnosis

End date: until the admitted patient recovered from DKA or discharged

### Data collection procedure and tool

#### Data collection tool

A structured data extraction tool was adapted by considering study variables such as socio-demographic, biochemical, clinical, and treatment predictors from literature and also by using WHO DKA follow sheet forms. A pretest was done on 24 participants of the study in Debre Markose hospital.

#### Data collection procedure

Patient records were retrieved using their unique registration number identified in the registration books. After obtaining a permission letter from the concerned body, charts and records were reviewed and data were extracted using an English data extraction form by trained two BSC nurses and supervised by one BSC nurse. Only index cases were included and recurrent episodes of DKA were excluded to avoid selection bias.

#### Quality Assurance

To keep data quality the supervisor and data collectors were trained on how and what the information they should collect from the targeted data sources. Data extraction forms were checked for completeness and missing information before data collection. The completeness of the collected data was checked onsite daily during data collection and give prompt feedback by the supervisor. Besides this, the principal investigator was carefully entered and thoroughly cleans the data before the commencement of the analysis.

#### Data Processing and Analysis

The data were coded, cleaned, and checked for completeness and consistency. Data were entered into Epi-data version 4.6 and analyzed using Stata Version 14.2.0 software. A Kaplan Meier survival curve was used to estimate DKA-free survival time. In addition, a generalized log-rank test was utilized to compare DKA free survival time between different categorical explanatory variables. Pair-wise comparison tests were done to check the presence of multicollinearity between each independent variable. Bivariable and Multivariable Cox proportional hazards models were fitted to identify predictors of time to DKA recovery incidence. The necessary assumptions of the Cox proportional hazards model were verified by log minus log plot, COX-Snell residuals for the goodness of fit and global test. Variables with P value < 0.25 in Bivariable analysis were entered into the multivariable Cox proportional hazards model to identify predictors of recovery time at p≤ 0.05(45). Hours were used as a time scale to calculate time to recovery. The outcome of each participant was dichotomized into censored or event (recovered).

#### Ethical Consideration

Ethical clearance was obtained from the institutional review board of Bahir Dar University, College of Medicine and Health Sciences (Ref. No: MD/11724/144). The formal letter was submitted to Debre-Markos referrals hospital for data collection and permission was assured. All information collected from patient cards was kept strictly confidential. Consent was not requested as it was a retrospective study.

## Result

### Socio-demographic characteristics

A total of 1281 adult DKA patients were admitted to Debre Markos referral hospital during the study period. Four hundred sixty -four (464) patients’ charts were selected by computer-generated random sampling technique and 452 patient’s charts were eligible among selected charts for this study and were included in the study. The median age of the participants was 25 years IQR (20-33). The majority of DKA adult patients were males and came from rural areas 295(65.27%) and 287(63.50%) respectively (Table 2).

**Table 1:**
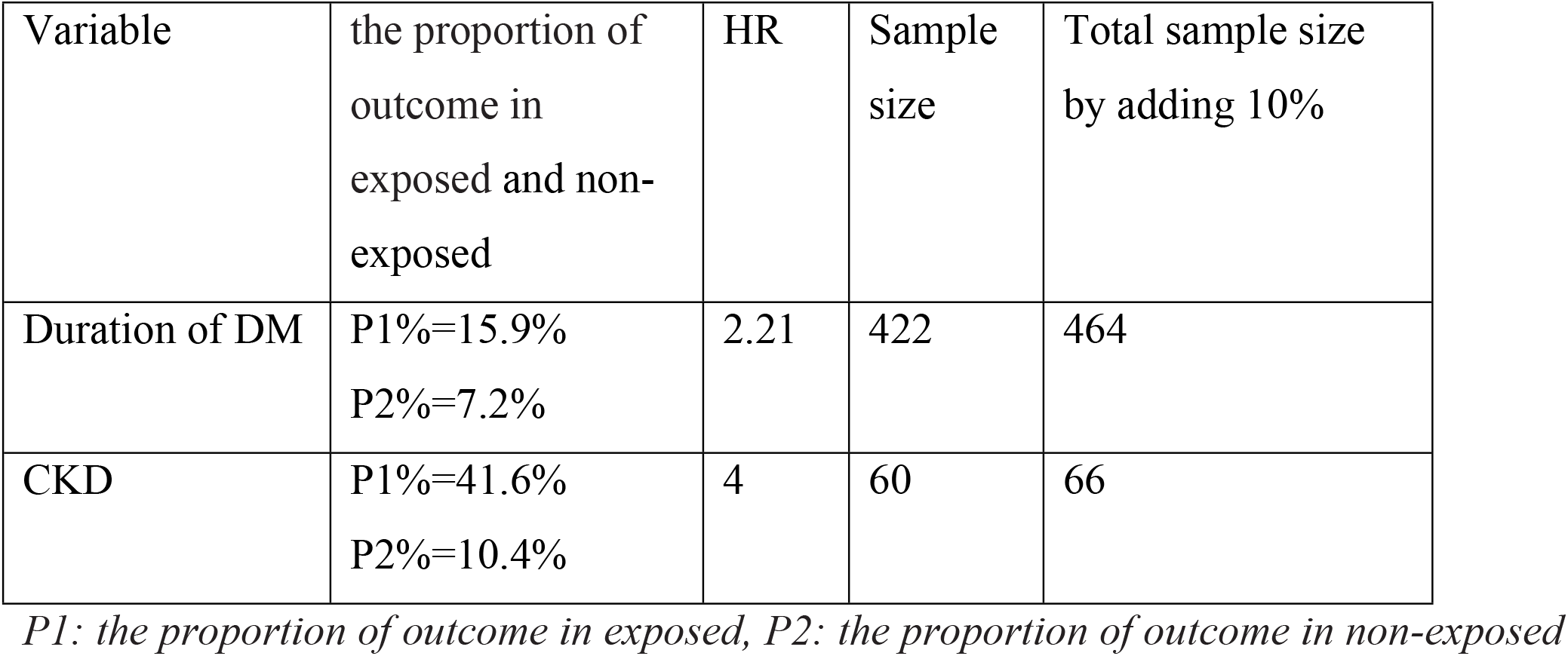
Sample size calculation for predictors of recovery of DKA among adults in Deber Markose Referral Hospital, North West Ethiopia 2021

**Table 2:** Socio-demographic characteristics among adult DKA patients, DMRH, North West Ethiopia, January 1, 2016 to January 1, 2021

| Variable | Category | Frequency | Percent (%) |
| --- | --- | --- | --- |
| Age(years) | 15-24 | 216 | 47.8 |
|  | 25-34 | 132 | 29.2 |
|  | 35-44 | 49 | 10.84 |
|  | 45-54 | 39 | 8.63 |
| | $\geq 55$ | 16 | 3.54 |
| Sex | Male | 295) | 65.3 |
|  | Female | 157 | 34.7 |
| Residence | Urban | 165 | 36.5 |
|  | Rural | 287 | 63.5 |

### Clinical characteristics and biochemical profile

Most DKA adult patients 296(65.49%) had previous DM history and 376 (83.19%) were T1DM. The most common clinical presentations of adult DKA patients were poly symptoms (97.35%), nausea/vomiting (64.30%), and fatigue (53.10%). The median time of duration of DKA symptoms till the treatment of the participant was 48hr IQR (96-24). Adult DKA patient’s clinical presentation of the severity of DKA mild, moderate, and severe was 64.38%, 22.57%, and 13.05% respectively. One hundred ninety-six (43.36%) of the study participants had previous DKA episodes, also 72(15.93%) and 159(35.18%) had an infection before DKA and acute recent illness respectively.

Among adult DKA patients, who had a DM history 89(19.69%) had co-morbidity. The most frequent comorbidity was HTN 40 (44.94%) (Table3).

**Table 3:**
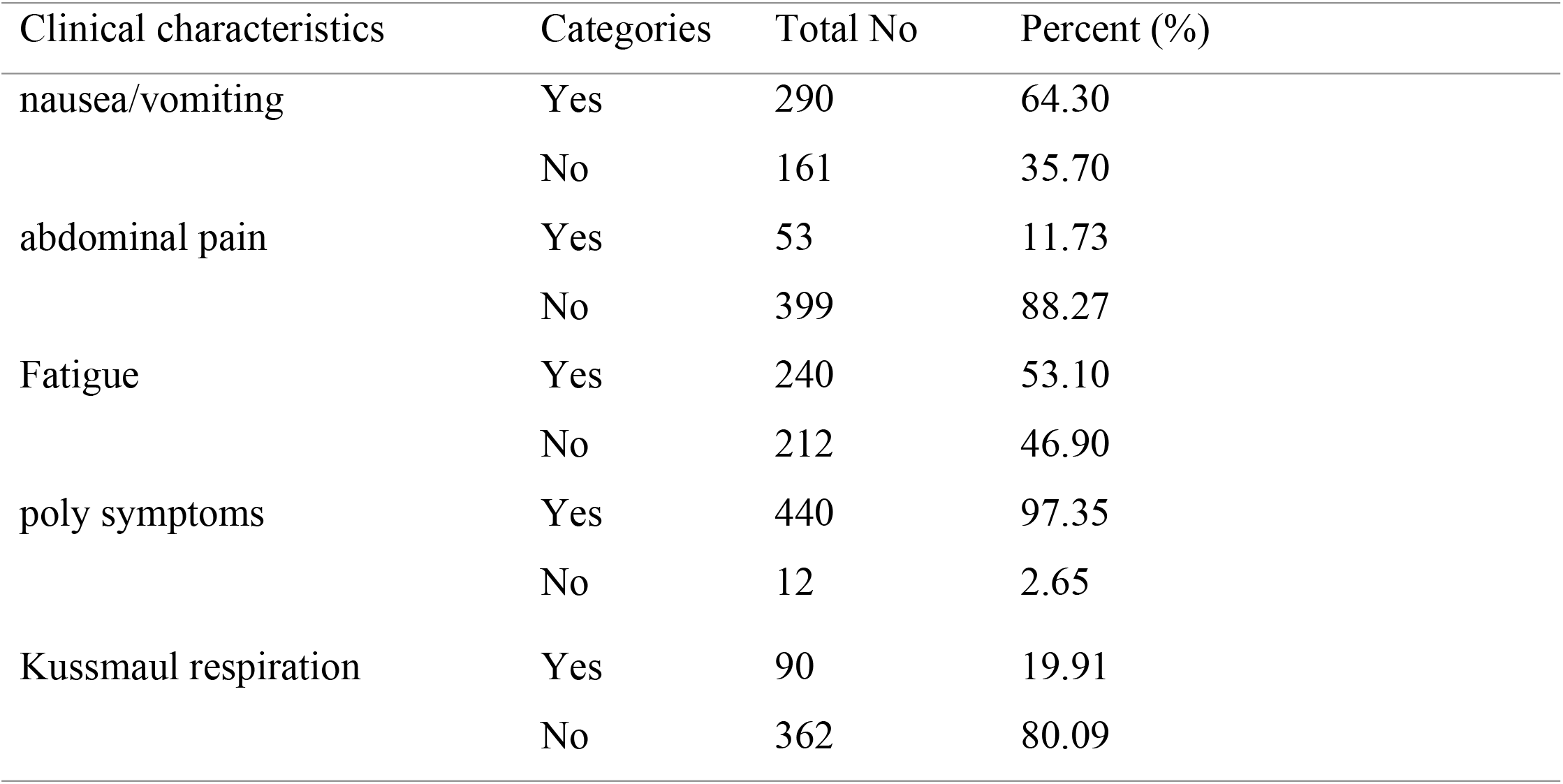
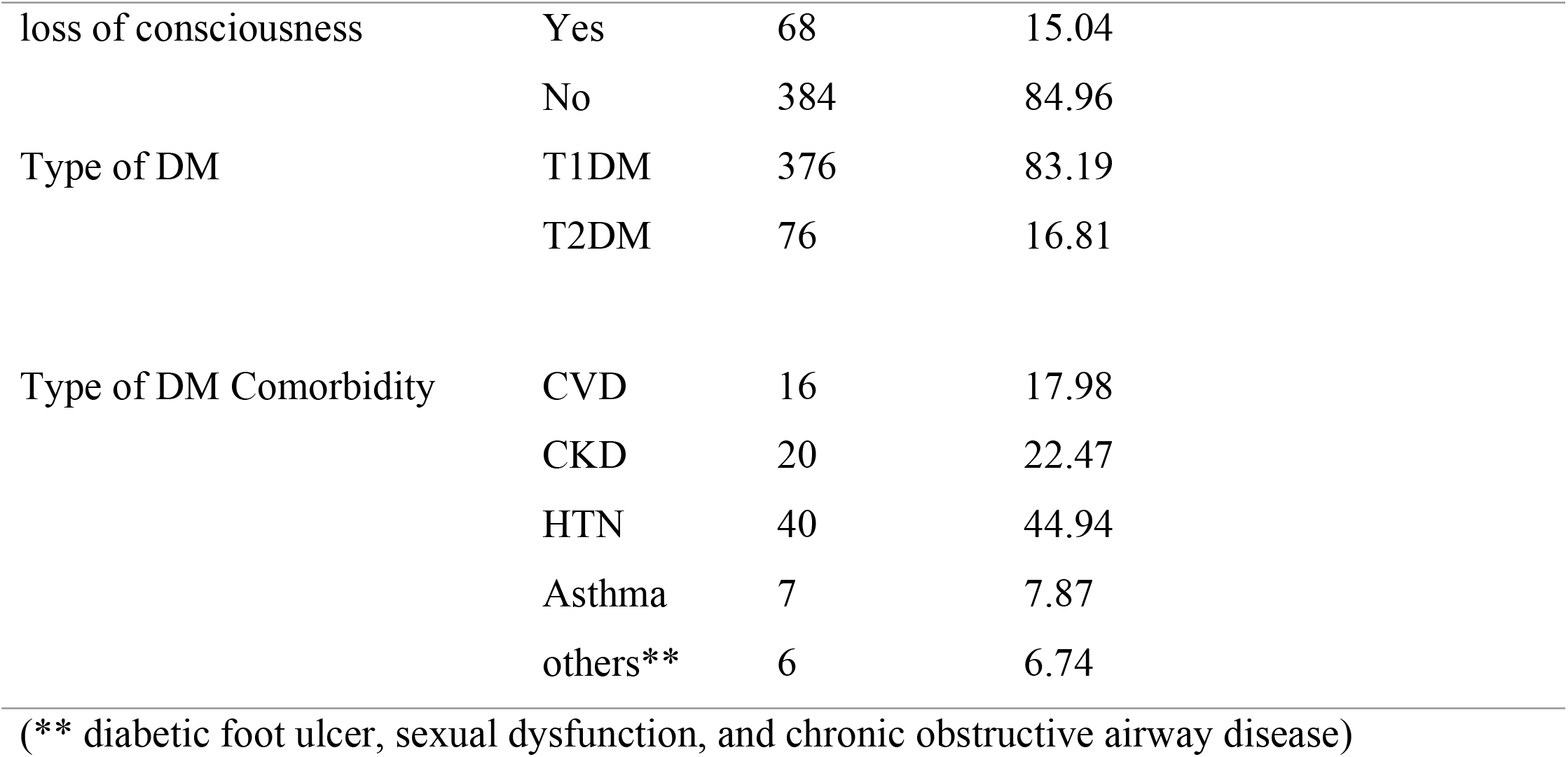
Baseline clinical characteristics among adult DKA patients, DMRH, North West Ethiopia, January 1, 2016 to January 1, 2021

| Clinical characteristics | Categories | Total No | Percent (%) |
| --- | --- | --- | --- |
| nausea/vomiting | Yes | 290 | 64.30 |
|  | No | 161 | 35.70 |
| abdominal pain | Yes | 53 | 11.73 |
|  | No | 399 | 88.27 |
| Fatigue | Yes | 240 | 53.10 |
|  | No | 212 | 46.90 |
| poly symptoms | Yes | 440 | 97.35 |
|  | No | 12 | 2.65 |
| Kussmaul respiration | Yes | 90 | 19.91 |
|  | No | 362 | 80.09 |
| loss of consciousness | Yes | 68 | 15.04 |
|  | No | 384 | 84.96 |
| Type of DM | T1DM | 376 | 83.19 |
|  | T2DM | 76 | 16.81 |
| Type of DM Comorbidity | CVD | 16 | 17.98 |
|  | CKD | 20 | 22.47 |
|  | HTN | 40 | 44.94 |
|  | Asthma | 7 | 7.87 |
|  | others** | 6 | 6.74 |

(\*\* diabetic foot ulcer, sexual dysfunction, and chronic obstructive airway disease)

### Time to recovery of adult DKA patients

The median time to recovery from DKA for all observations was 24 hrs. IQR (12-37). The median recovery time of adults from DKA was different regarding different categories of biochemical and clinical characteristics. For instance, the median recovery time of mild DKA and moderate DKA was 18hrs and 30 hrs. Whereas in severe DKA was 49hrs (Table 4).

**Table 4:** The median time to recovery, and comparison of DKA free time among adult DKA patients, DMRH, North West Ethiopia, January 1, 2016 to January 1, 2021

| Variable | Category | Survival status of DKA |  | Total No (%) | Median recovery time (95%CI) | Log-rank (P-value) |
| --- | --- | --- | --- | --- | --- | --- |
|  |  | Recovered No (%) | censored No (%) |  |  |  |
| DM history | Yes | 282(65.73) | 14(60.87) | 296(65.49) | 24(23-25) | 0.73 |
|  | NO | 147(34.27) | 9(39.13) | 156(34.51) | 24(22-27) |  |
| Type of DM | T1DM | 355 (82.75) | 21(91.30) | 376(83.19) | 24(23-25) | 0.62 |
|  | T2DM | 74 (17.25) | 2(8.70) | 76(16.81) | 25(20-28) |  |
| Diabetes duration | ≤ 5 years | 210(48.95) | 10(43.48) | 219(48.67) | 28(25 -30) | <0.001* |
|  | >5 years | 109(25.41) | 9(39.13) | 118(26.11) | 16(14 -20) |  |
|  | Newly diagnosis | 110(25.64) | 4(17.39) | 114(25.22) | 24(18-25) |  |
| Comorbidity of DM | Yes | 86(20.05) | 3(13.04) | 89(19.69) | 26(24 -30) | 0.31 |
|  | NO | 343(79.95) | 20(86.96) | 363(80.31) | 24(23-25) |  |
| history of DKA episodes | Yes | 188(43.82) | 8(34.78) | 196(43.36) | 26(24 -28) | 0.04* |
|  | NO | 241(56.18) | 15(65.22) | 256(56.64) | 24(20-24) |  |
| Severity of DKA | Mild | 278(64.80 ) | 13(56.52) | 291(64.38) | 18(15-20) | <0.001* |
|  | Moderate | 96 (22.38) | 6(26.09) | 102(22.57) | 30(26-40) |  |
|  | Severe | 55(12.82) | 4(17.39) | 59(13.05) | 49(39 -72) |  |
| Duration of DKA | <=24 hrs. | 150 (34.97) | 7(30.43) | 153(33.85) | 20(15-24) | <0.001* |
|  | 25-48hrs | 75(17.48) | 5(21.74) | 74(16.37) | 24(19 -26) |  |
|  | 49-96hrs | 143(33.33) | 7(30.43) | 142(31.42) | 26(25-29) |  |
|  | ≥97hrs | 61(14.22) | 4(17.39) | 83(18.36) | 25(23-33) |  |
| infection before DKA | Yes | 71(16.55) | 1(4.35) | 72(15.93) | 30(25-34) | 0.005* |
|  | NO | 358(83.45) | 22(95.65) | 380(84.07) | 24(22-24) |  |
| acute recent illness | Yes | 149(34.73) | 10(43.48) | 159(35.18) | 30(26-35) | <0.001* |
|  | NO | 280(65.27) | 13(56.52) | 293(64.82) | 22(19- 24) |  |
| RBS in mg/dL | ≤500 | 254(59.21) | 14(60.87) | 268(59.29) | 20(17-23) | <0.001* |
|  | >500 | 175(40.79 ) | 9(39.13) | 184(40.71) | 31(27 -35) |  |

### The incidence rate of recovery from DKA

From 452 study participants, 429 (94.9%) adults were recovered and the rest 23(5.1%) were censored observations. The lowest and the highest length of follow-up were 3.5 and 102 hours respectively, and the total person-time risk was 12546.513. The overall recovery rate from DKA was 3.42 per 100 person-hours observation (95% CI: 3.11–3.76). The recovery rate among severity of DKA in mild, moderate, and severe was 4.93 per 100 person-hours (95% CI: 4.38-5.54), 2.61 per 100 person-hours (95% CI: 2.14 -3.19), and 1.71 per 100 person-hours (95% CI: 1.31–2.22) respectively.

### Survival estimates for time to recovery

The DKA free time of adults with DKA was estimated by the Kaplan-Meier survival curve. The curve tends to decrease rapidly within the first 24hrs indicating that most adults recovered from DKA within this time (Fig 1). The survival estimates of DKA patients were varied among severity, duration of DM, duration of DKA, and RBS level (Figs 2-4).

**Figure 1:**
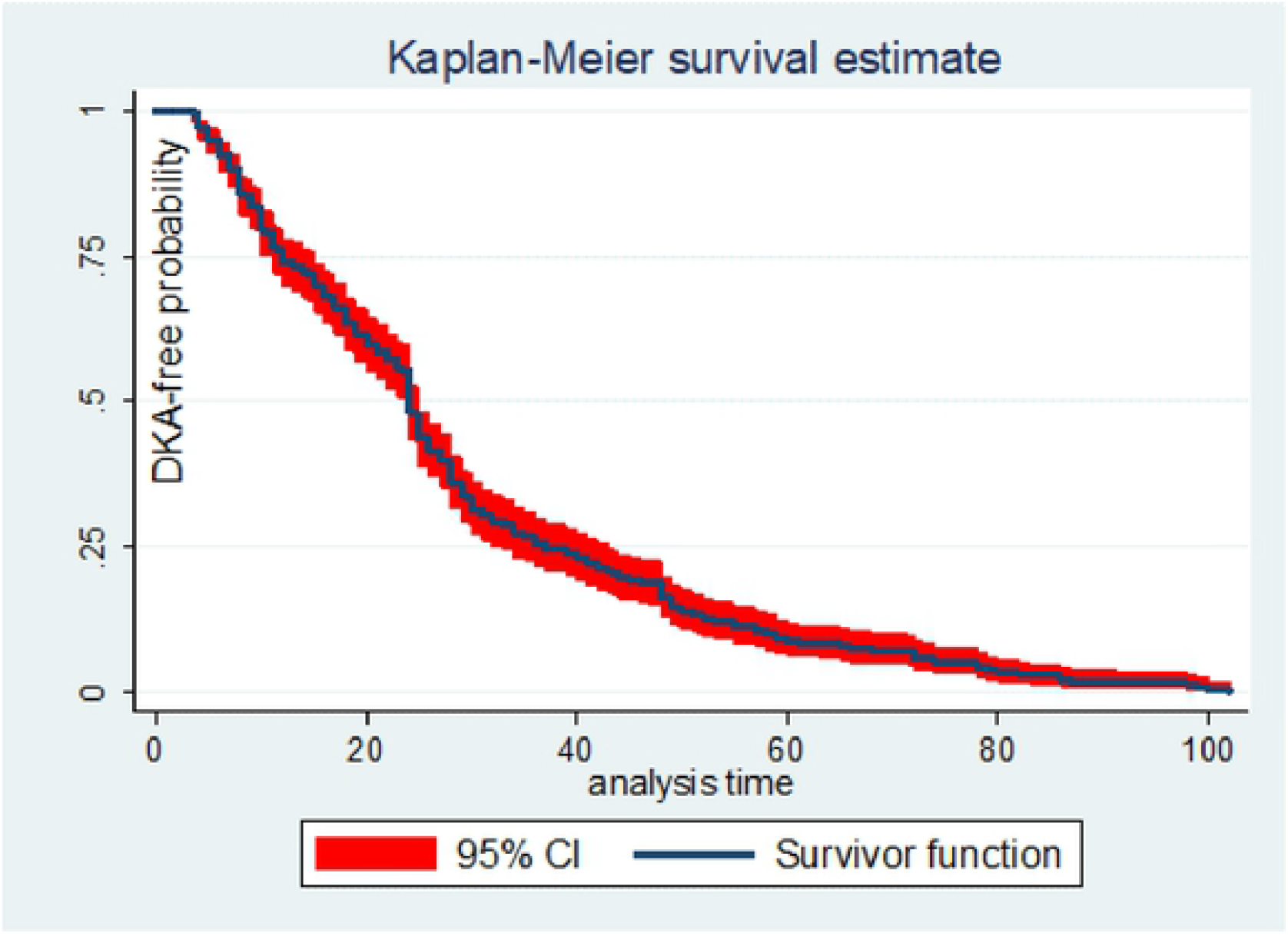
Overall Kaplan -Meier estimation of DKA free time functions of patients on adult DKA patients, DMRH, North West Ethiopia, January 1 2016 to January 1, 2021

**Figure 2:**
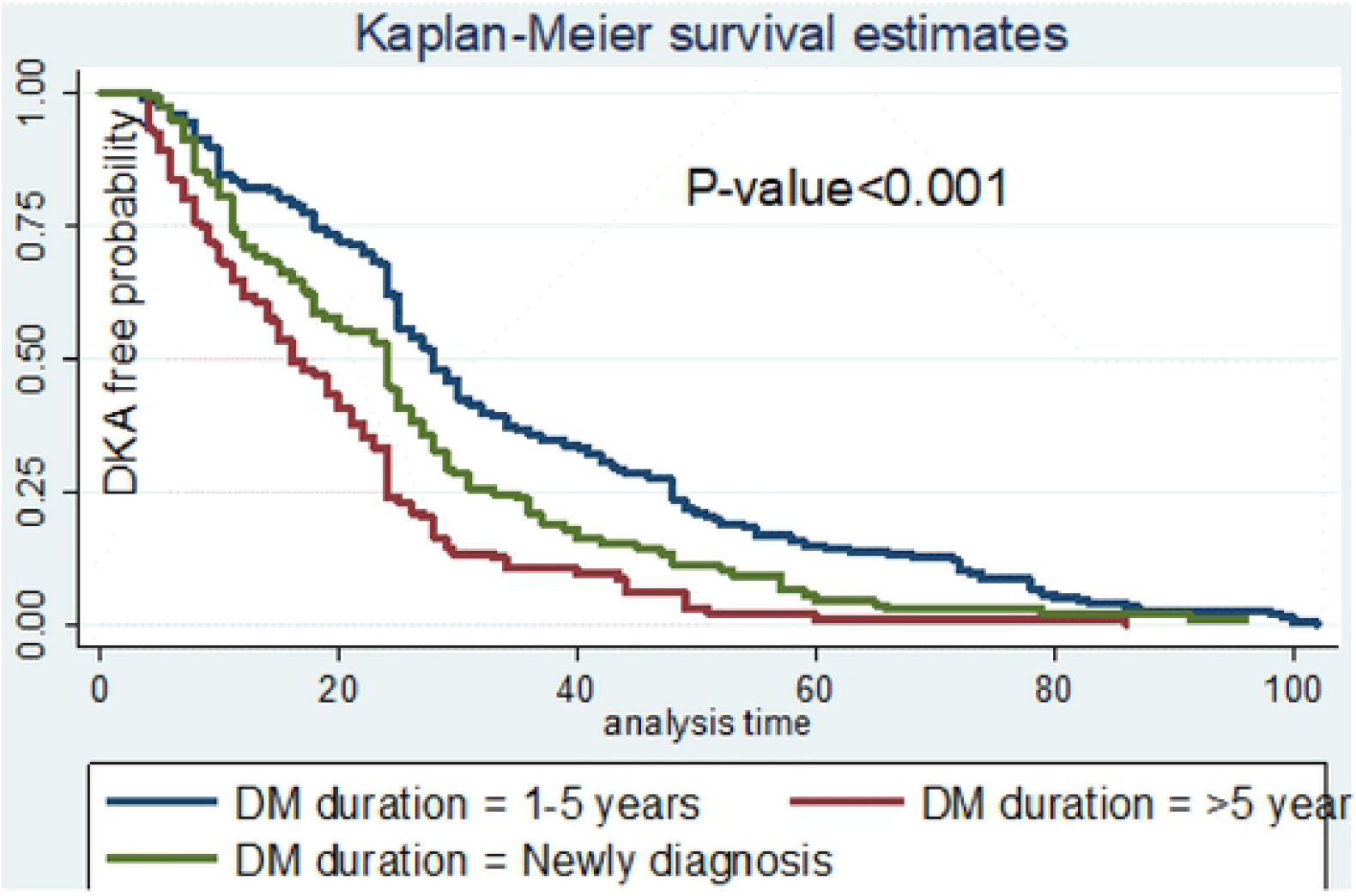
The Kaplan-Meier survival curves compare DKA free time of adult DKA patients between duration of DM, DMRH, North West Ethiopia, January 1 2016 to January 1 2021

**Figure 3:**
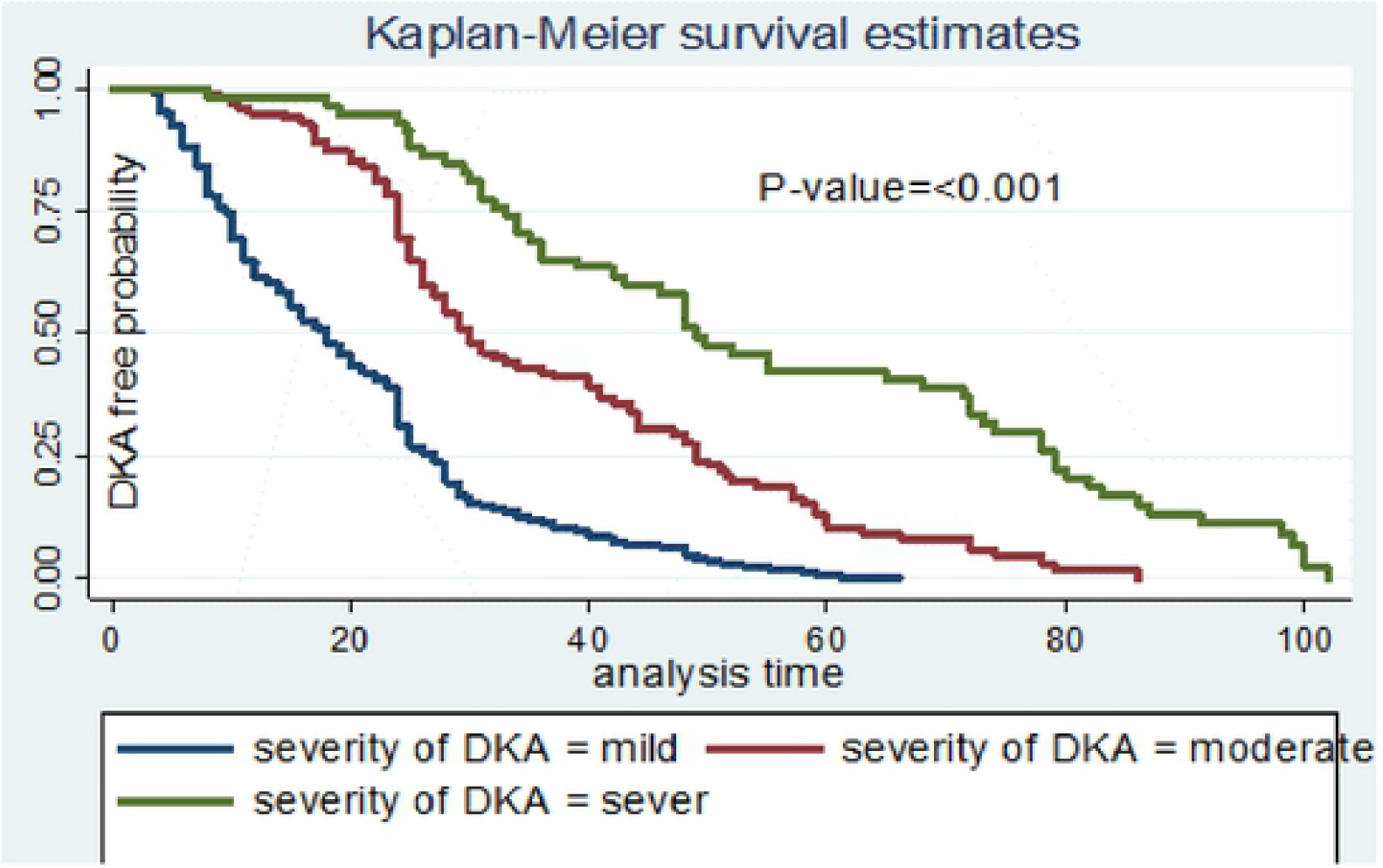
The Kaplan-Meier survival curves DKA free time of adult DKA patients between severity of DKA, DMRH, North West Ethiopia, January 1 2016 to January 1, 2021

**Figure 4:**
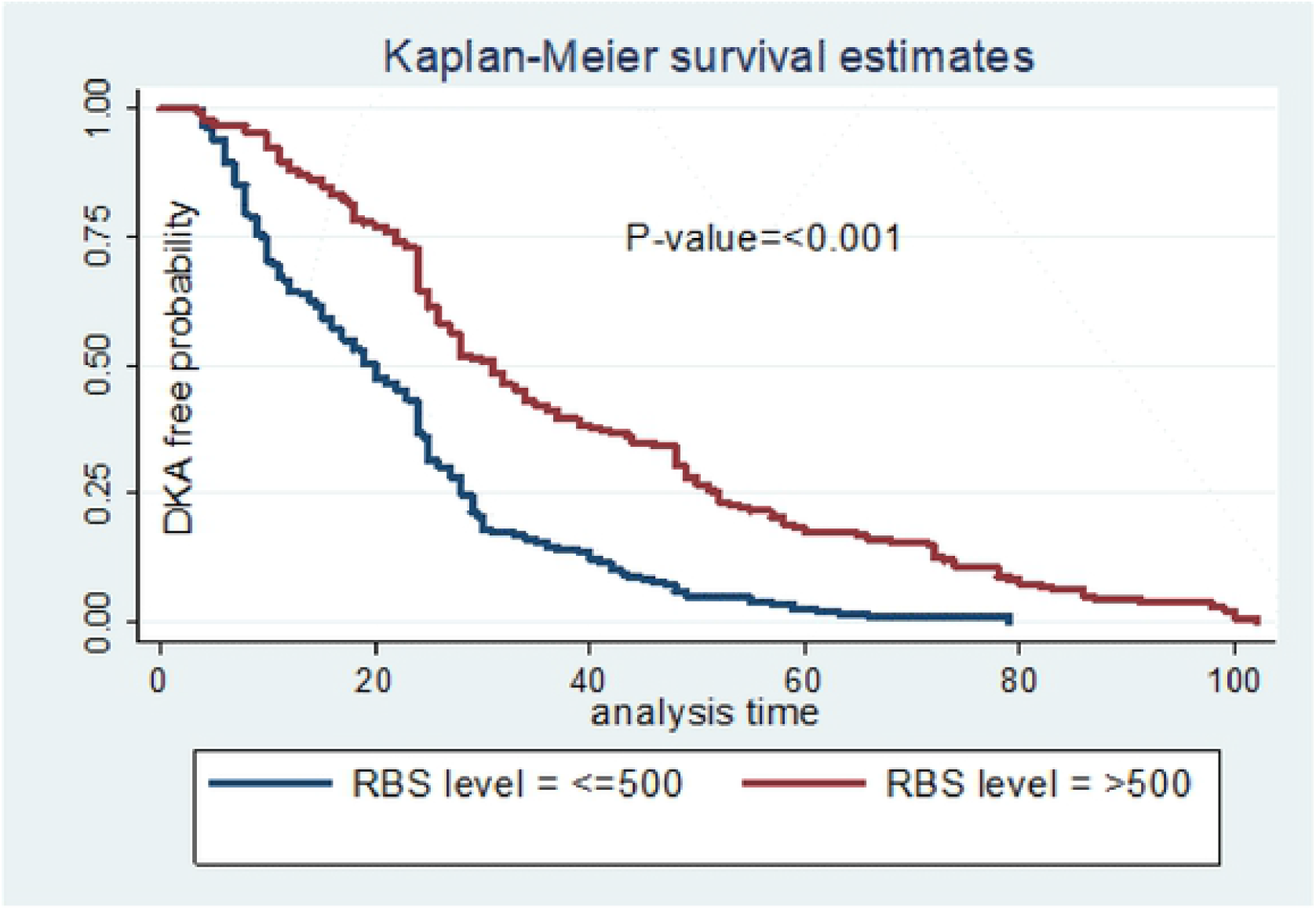
The Kaplan-Meier survival curves compare DKA free time of adult DKA patients between RBS level, DMRH, North West Ethiopia, January 1 2016 to January 1, 2021

### Comparison of survival status

Log-Rank test was used to compare survival time between categories of different predictors. Based on this test, survival time among different groups of predictors such as severity, duration of DM, duration of DKA, presence of infection before DKA and acute recent illness, and RBS level were significantly different in survival time at a 5% level of significance (Tables 2 & 3).

### Predictors of recovery time from DKA

In the Bivariable Cox, Proportional Hazards regression model; DM duration, infection before DKA, duration of DKA, acute recent illness, RBS level, and severity of DKA were all associated with DKA recovery (P <0.05). Predictors that had association at a p-value of ≤ 0.25 in bivariable Cox regression and non-collinear independent variables were included in multivariable Cox regression. In the multivariable analysis, the AHR of the severity of DKA, duration of DKA, DM duration, and RBS level was estimated as independent predictors of time to recovery with a value of p<0.05.

The recovery time was delayed by 76% (AHR=0.24, 95% CI=0.16-0.35) among severe DKA adult patients as compared with mild DKA adult patients. Also, the recovery time was delayed by 54% (AHR=0.46, 95%CI=0.35-0.59) among moderate DKA adult patients as compared with mild DKA adult patients. Adult DKA patients with RBS >500mg/dL had slower recovery as compared with those who had RBS ≤500mg/dL (AHR=0.64, 95%CI= (0.51-0.79). DKA adult patients with >5years duration of DM follow-up had 74% faster recovery as compared with those who had a follow-up of DM duration ≤ 5 years (AHR=1.74, 95%CI 1.35-2.25).

Adult DKA patients with ≥97hrs of duration DKA till treatment had slower recovery as compared with those who had ≤24hrs (AHR=0.46, 95%CI 0.33-0.64).

### Overall model fitness test

Figure (5) below shows the overall model fitness of the data in Cox Proportional hazards regression model. In the present finding, the hazard function follows the 45° line very closely. This indicates as the Cox model does fit these data to reasonable. Hence, the Cox Snell residuals test shows overall the goodness of fitness of the model (the model is a good fit).

**Figure 5:**
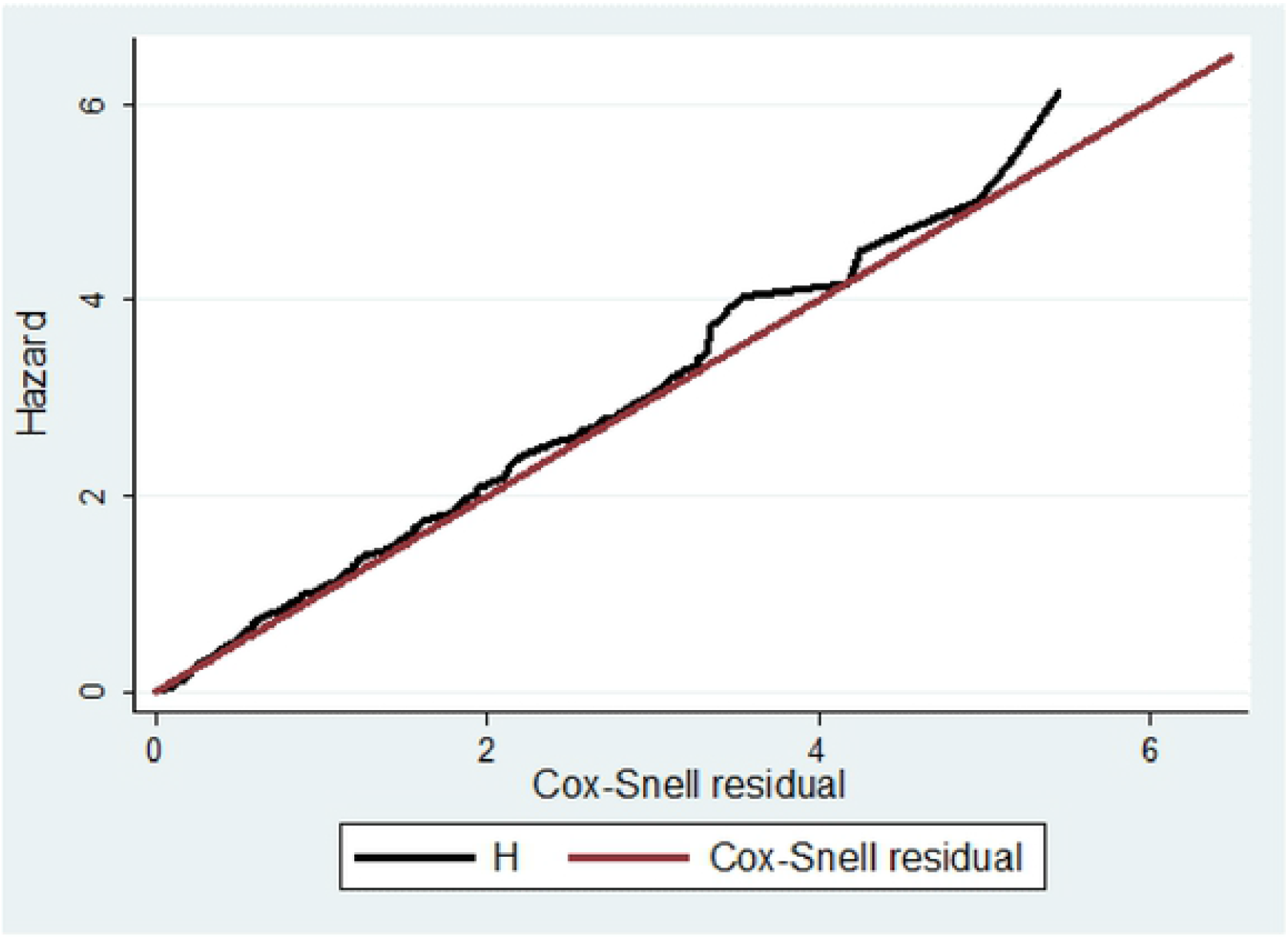
Cox Snell residual test showing overall goodness of fit of the Cox proportional hazards model.

### Test of proportional-hazards assumption

The findings indicated that all variables included in the model satisfied PH assumptions (p value>0.05). So the model is a good fit (Table 6).

**Table 5:-.**
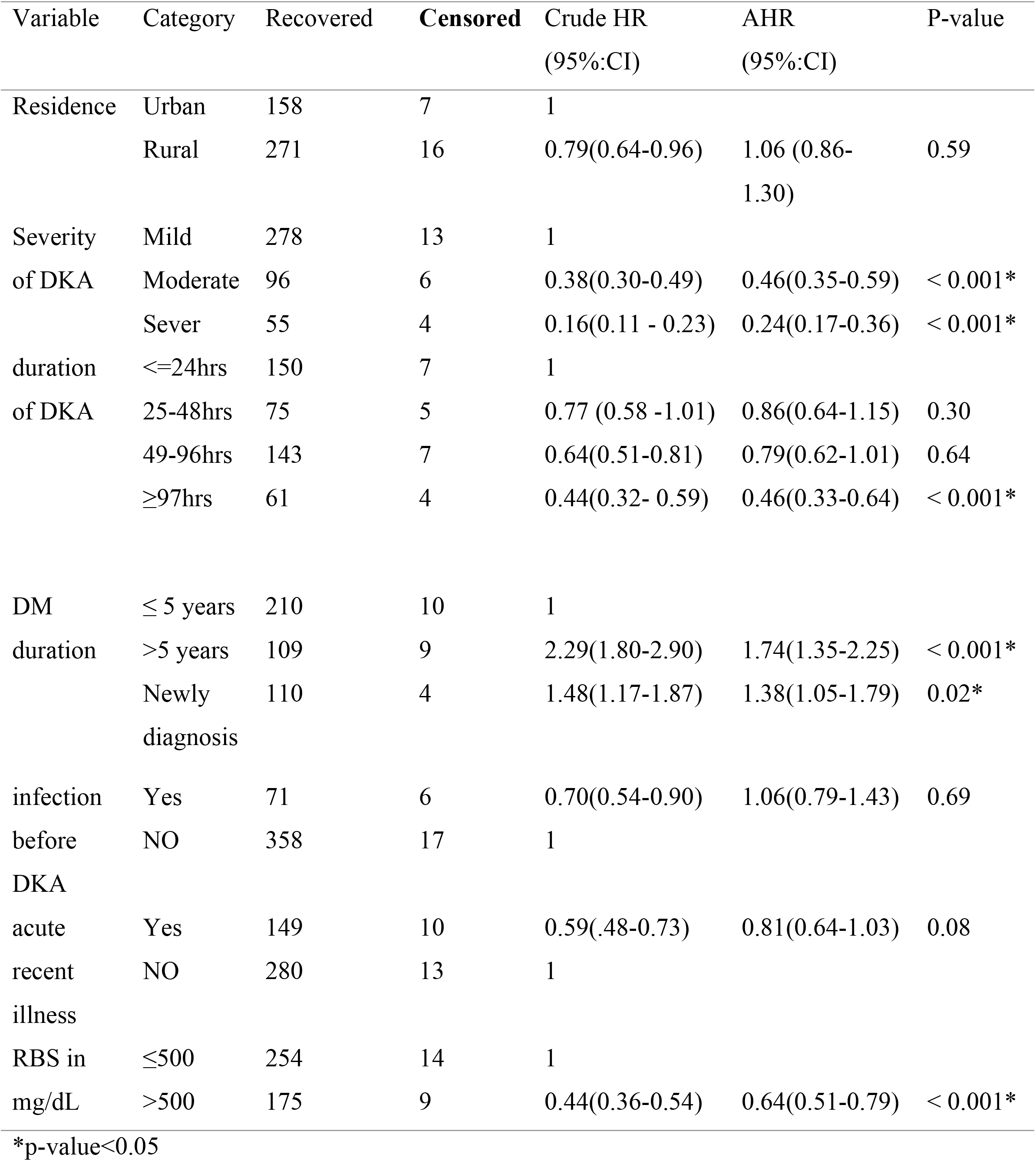
Results of the Bivariable and Multivariable cox proportional hazards regression analysis of adult DKA patients, DMRH, North West Ethiopia, January 1 2016 to January 1 2021

**Table 6:-.**
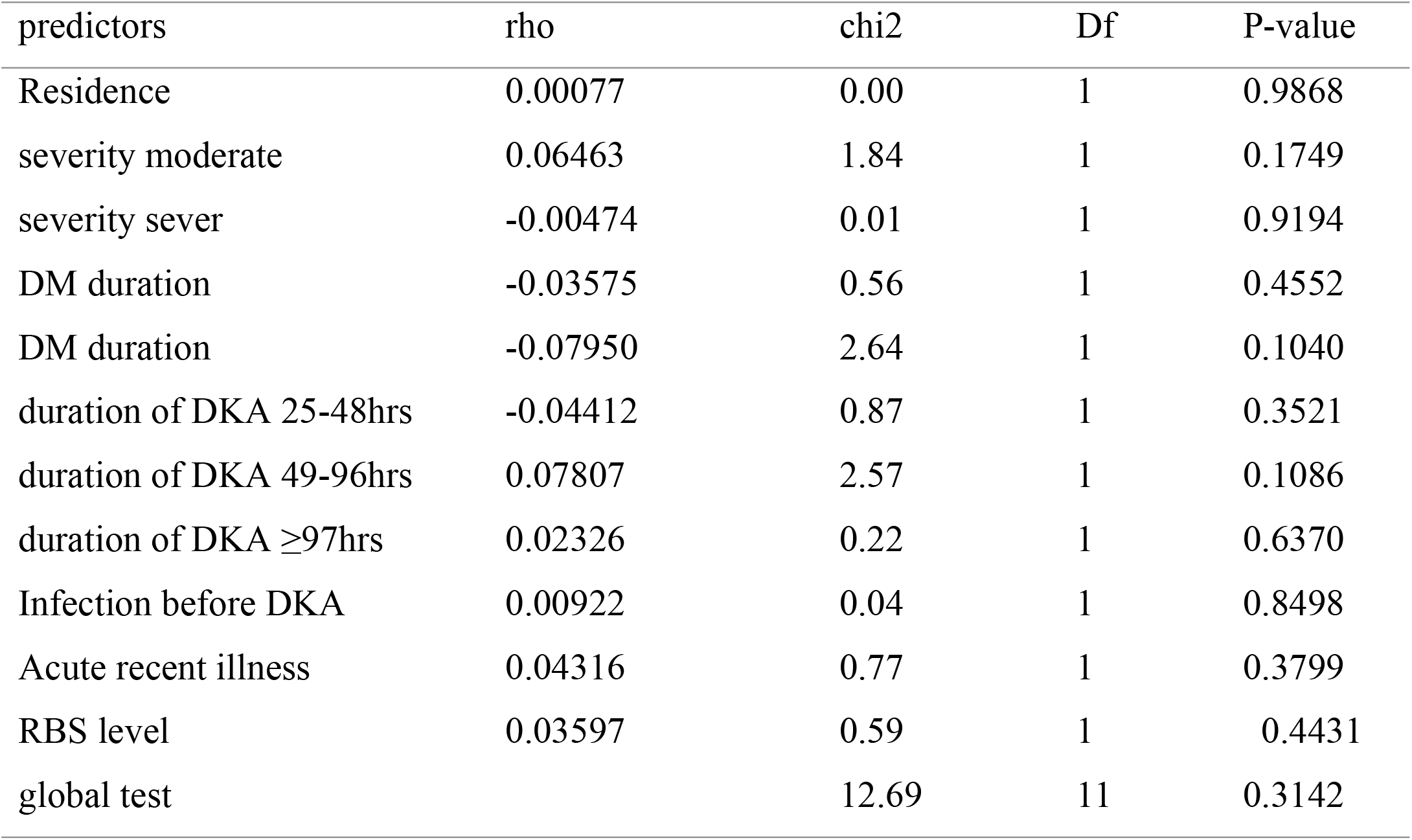
Goodness-of-fit test is assessing proportional hazards Assumption

| predictors | rho | chi2 | Df | P-value |
| --- | --- | --- | --- | --- |
| Residence | 0.00077 | 0.00 | 1 | 0.9868 |
| severity moderate | 0.06463 | 1.84 | 1 | 0.1749 |
| severity sever | -0.00474 | 0.01 | 1 | 0.9194 |
| DM duration | -0.03575 | 0.56 | 1 | 0.4552 |
| DM duration | -0.07950 | 2.64 | 1 | 0.1040 |
| duration of DKA 25-48hrs | -0.04412 | 0.87 | 1 | 0.3521 |
| duration of DKA 49-96hrs | 0.07807 | 2.57 | 1 | 0.1086 |
| duration of DKA ≥97hrs | 0.02326 | 0.22 | 1 | 0.6370 |
| Infection before DKA | 0.00922 | 0.04 | 1 | 0.8498 |
| Acute recent illness | 0.04316 | 0.77 | 1 | 0.3799 |
| RBS level | 0.03597 | 0.59 | 1 | 0.4431 |
| global test |  | 12.69 | 11 | 0.3142 |

## Discussion

This study was conducted to determine the time to recovery from DKA and its predictors. The median time to recovery was 24hrs (IQR 12-37). The finding of this study is almost consistent with what was observed in a study conducted in South Africa(28), and prolonged when compared with studies conducted in Australia(30) and in Thailand(31).On the contrary, it is faster than a study done in Kenya(29). The possible reason for this discrepancy could be related to the difference in the standard treatment protocol because studies show that there is an improved clinical outcome associated with insulin pump therapy compared with injection therapy(9, 46). The other possible reason may be due to low socio-economic status since low educational level increased the risk of severe DKA that leads to prolonged recovery time (47).

The severity of DKA, duration of DKA, DM duration, and RBS level were independent predictors of time to recovery. The recovery time was delayed by 76% among severe DKA adult patients as compared with mild DKA adult patients. This finding is supported by the study conducted in South Africa identifies severe DKA was correlated with a longer time to resolution (28). The possible reason in this study could be due to the disease pathophysiology, patients with severe DKA had more electrolyte abnormalities compared with the mild and moderate forms of the disease(48) which is supported with a study conducted in Australia showed that higher admission potassium levels are independent predictors of a slower time to resolution of DKA(30).

Adult DKA patients with RBS >500mg/dL had slower recovery as compared with those who had RBS ≤500mg/dL. This finding is supported by a study that revealed initial blood glucose was a significant predictor of rapid resolution of DKA patients(36). This is possibly due to extremely high blood glucose concentrations result in loss of blood volume, low blood pressure, and impaired central nervous system function (hyperglycemic coma) that prolonged DKA recovery (49).

DKA adult patients with >5years duration of DM follow-up had 74% faster recovery as compared with those who had follow-up DM duration ≤ 5 years. This finding is consistent with a study in Saudi Arabia, patients who had a duration of one to five years of diabetes mellitus were almost five times more likely to get out of DKA in more than 72 hours when compared with those who had a duration of more than five years(33). A shorter duration of diabetes has been associated with both higher levels of HbA_1c_ and risk of all-cause hospitalizations in adults (50). In this study, a shorter duration of diabetes was predictive of slower recovery of diabetic ketoacidosis. This may be due to those DKA patients with a shorter duration of diabetes may be less knowledgeable and experienced with diabetes management, and correspondingly less adherent to insulin adjustment and diabetes management(51).

Adult DKA patients with ≥97hrs of duration DKA till treatment had slower recovery as compared with those who had ≤24 hrs. This finding is in line with a study performed in Kenya that revealed delays in initiation of management have contributed to the prolonged DKA resolution times (29). This could be due to timely diagnosis and initiation of management of DKA is associated with faster recovery(9).

## Conclusions

The median time to recovery from DKA was relatively prolonged. The severity of DKA, duration of DKA, DM duration, and RBS level were statistically significant predictors of time to recovery time from DKA. Therefore, we highly recommend Health care providers shall closely monitor and follow particularly for those who have a shorter duration of DM follow-up and severe DKA and give health education to the DM patients to come the health facility immediately when they become ill. We also recommend further study using a prospective design by including admission pH and admission serum potassium level to fill its limitation.

## Data Availability

All relevant data are within the manuscript and its Supporting Information files

## Abbreviations and Acronyms

ADA: American Diabetes Association
BG: Blood Glucose
CKD: Chronic Kidney Disease
CVD: Cardio Vascular Disorder
DKA: Diabetic Ketoacidosis
DM: Diabetes Mellitus
ETB: Ethiopian Birr
HTN: Hypertension
IV: Intravenous
RBS: Random Blood Sugar
T1DM: Type 1 Diabetes Mellitus
T2DM: Type 2 Diabetes Mellitus
UK: United Kingdom
USA: United States of America

## Declarations

### Consent for publication

Not applicable

### Availability of data and material

All relevant data’s for this study presented within the manuscript and supportive files.

### Authors’ contributions

**DT**: Formulating research problem, design of study, data collection, analysis, interpretation, conclusion, preparing initial manuscript draft

**GD, YM, BBT-** Actively participated in data collection, analysis and interpretation, and writing up manuscript. All authors had thoroughly read and approved the manuscript.

## Acknowledgment

We would like to express our deepest gratitude to Bahir Dar University, College of Medicine and Health Science for letting us to conduct this. Next, we also advance our profound thanks to Debere Markose referral Hospital staff working at diabetes Clinic, Card room and administration position providing us valuable data’s on source population and aiding us on chart retrieval. Last but not least, our special thanks go to data collectors and the supervisor for their willingness, commitment, and intensive works during data collection for the betterment of this research.

